# Methylprednisolone as Adjunct to Endovascular Thrombectomy for Acute Ischemic Stroke Patients with a Large Infarct Core (MIRACLE): Protocol of a Randomized Trial

**DOI:** 10.1101/2025.10.03.25337303

**Authors:** Ying Fu, Liangliang Qiu, Qianqian Lin, Yi Lin, Wenlong Zhao, Kunxin Lin, Changwei Guo, Zhangbao Guo, Zhongming Qiu, Thanh N. Nguyen, Minting Lin, Duolao Wang, Ning Wang, Wenjie Zi, Wanjin Chen, the MIRACLE investigators

## Abstract

**Background:** Despite successful endovascular thrombectomy (EVT) for acute ischemic stroke (AIS) with a large infarct core, patient disability and mortality remain high.

**Aim:** The MIRACLE trial aims to evaluate the efficacy and safety of administering adjunct intravenous methylprednisolone in AIS patients with a large infarct core.

**Design:** Methylprednisolone as Adjunct to Endovascular Thrombectomy for patients with Acute Large Ischemic Stroke (MIRACLE) is a multicenter, prospective, randomized, double-blind, placebo-controlled phase 3 trial conducted across 99 stroke centers in China. A total of 902 patients with anterior circulation large vessel occlusion and large infarct core (Alberta Stroke Program Early CT Score [ASPECTS] <6 or infarct volume ≥50 mL on CT perfusion) presenting within 12 hours of last known well time will be randomized 1:1 to receive either intravenous methylprednisolone (2 mg/kg/day, max 160 mg) or its corresponding placebo for three consecutive days, starting immediately after randomisation.

**Study outcomes:** The primary efficacy outcome is all-cause mortality at 90 days. Key secondary outcomes include functional status assessed by the modified Rankin Scale (mRS), neurological status assessed by National Institutes of Health Stroke Scale (NIHSS), quantitative cerebral edema markers (midline shift, relative hemispheric volume, and water uptake), and decompressive craniectomy rates. The primary safety outcome is symptomatic intracranial hemorrhage (SICH) within 48 hours post-EVT. Analyses will follow the intention-to-treat principle. Recruitment began in August 2024 and is expected to conclude by October 2025.

**Conclusion:** The MIRACLE trial will provide evidence of the efficacy and safety of adjunctive methylprednisolone in AIS patients with a large core undergoing EVT within 12 hours of last known well.

**Trial Registration:** ClinicalTrials.gov NCT06360458

**CLINICAL PERSPECTIVE:** *What Is New?:* The MARVEL trial (Methylprednisolone as Adjunct to Endovascular Thrombectomy for Large Vessel Occlusion Stroke) [1] demonstrated a potential safety benefit and reduced mortality with adjunctive methylprednisolone in patients with anterior circulation large vessel occlusion stroke undergoing endovascular thrombectomy (EVT), but it excluded patients with very large infarct core (Alberta Stroke Program Early CT Score [ASPECTS] <3 or volume >50-70 mL), leaving its efficacy in this high-risk population unknown. This study aims to investigate whether adjunctive methylprednisolone improves survival in acute ischemic stroke (AIS) patients with large infarct core undergoing EVT (ASPECTS < 6 and/or core infarct volume > 50 ml).

*What Aare the Clinical Implications?:* If positive, MIRACLE will provide robust evidence for a simple, accessible, and inexpensive adjunctive therapy to EVT that could improve outcomes for AIS patients with large infarct core.

## INTRODUCTION AND RATIONALE

Endovascular thrombectomy (EVT) has revolutionized the treatment of acute ischemic stroke (AIS) due to large vessel occlusion (LVO) [2]. Six randomized trials demonstrated the benefit of EVT even in patients with large infarct core, defined by low Alberta Stroke Program Early CT Score (ASPECTS) ( <6) or ischemic core volumes ≥50 mL [3-8]. Despite high rates of successful recanalization (80%-94%) [9], half of these patients experience poor functional outcomes or death [10, 11]. This discrepancy between angiographic success and clinical benefit is often attributed to reperfusion injury characterized by hemorrhagic transformation and malignant cerebral edema [12].

The peak of cerebral edema occurs within days after stroke onset and is a major cause of neurological deterioration and death, particularly in patients with large infarcts [13, 14]. The underlying pathophysiology involves a potent inflammatory cascade and ischemia, which can also be exacerbated by reperfusion [15, 16]. Corticosteroids, with their anti-inflammatory, immunomodulatory, and blood-brain barrier-stabilizing properties, have been proposed as potential neuroprotective agents to mitigate this process [17]. Preclinical studies demonstrated that early corticosteroid administration reduced brain edema, limited infarct expansion, and improved neurological outcomes in animal models of ischemic stroke [18, 19].

The Methylprednisolone as Adjunct to Endovascular Thrombectomy for Large Vessel Occlusion Stroke (MARVEL) trial evaluated methylprednisolone as an adjunct to EVT in a broad population of LVO stroke patients (ASPECTS 3–10) [1]. Although the primary outcome (ordinal shift in modified Rankin Scale [mRS] at 90 days) was not met, exploratory analyses suggested a potential reduction in mortality and symptomatic intracranial hemorrhage (SICH), particularly in the subgroup of patients with an intracranial carotid artery occlusion (ICA) [20]. However, patients with very large infarct core (ASPECTS <3 or core volume >50–70 mL)—those most vulnerable to life-threatening edema and haemorrhage—were excluded from the MARVEL trial. Thus, whether methylprednisolone confers an additional benefit in patients with large infarct core remains unknown.

The Methylprednisolone as Adjunct to Endovascular Thrombectomy for patients with Acute Large Ischemic Stroke (MIRACLE) trial was designed to address this knowledge gap by evaluating whether early administration of intravenous methylprednisolone following EVT improves survival and functional outcomes in AIS patients with large infarct core.

## METHODS

### Hypothesis

Early administration of intravenous methylprednisolone as an adjunct to EVT can reduce 90-day all-cause mortality as compared with EVT alone in acute ischemic stroke patients with large infarct core.

### Design

MIRACLE is an investigator-initiated, multicenter, prospective, randomized, double-blind, placebo-controlled phase 3 trial (Figure 1). The trial investigates intravenous methylprednisolone in addition to EVT in the interventional arm compared to EVT alone in the control group. The trial was registered at ClinicalTrials.gov (NCT06360458). The trial was designed in compliance with the Declaration of Helsinki. The study protocol has been approved by the central ethics committee at the First Affiliated Hospital of Fujian Medical University and all participating sites. This study is conducted at 99 stroke centers in China.

**Figure 1.**
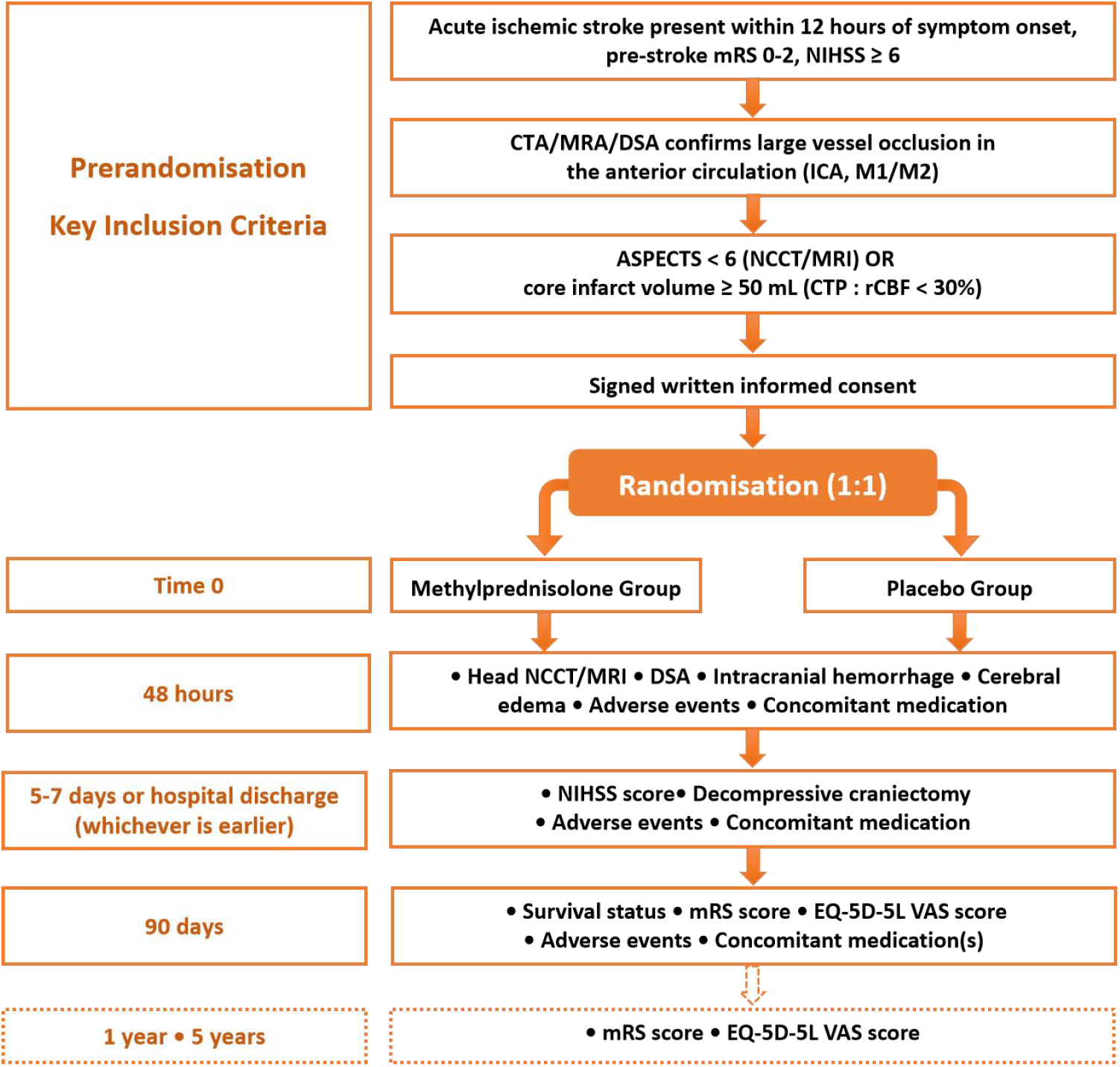
Study design: randomisation algorithm. ASPECTS, Alberta Stroke Program Early CT Score; CT, computed tomography; CTA, computed tomography angiography; CTP, CT perfusion; DSA, digital subtraction angiography; EQ-5D-5L VAS, European Quality Five-Dimension Five-Level Visual Analogue Scale; ICA, internal carotid artery; MCA, middle cerebral artery; MRA, magnetic resonance angiography; MRI, magnetic resonance imaging; mRS, modified Rankin Scale; NCCT, non-contrast CT; NIHSS, National Institutes of Health Stroke Scale.

### Eligibility criteria

The inclusion criteria for the trial consists of patients who present with symptoms of an acute ischemic stroke within 12 hours of symptom onset from time last known well, with National Institutes of Health Stroke Scale 6 or greater, baseline ASPECTS < 6 and/or core infarct volume > 50 ml, an occlusion of an intracranial internal carotid artery or middle cerebral artery (M1 or M2 segment), and planned for EVT. Patients are excluded if they have an allergy to glucocorticoid, known systemic infectious disease, or mRS > 2 prior to stroke onset. Box 1 lists the complete inclusion and exclusion criteria.

#### Box 1

##### Inclusion criteria

1. Age ≥18 years;
2. The time from last known well to randomization is within 12 hours;
3. Anterior circulation ischemic stroke is according to clinical symptoms or imaging examination;
4. Baseline National Institutes of Health Stroke Scale (NIHSS) ≥6;
5. Baseline ASPECTS <6 (based on non-contrast CT [NCCT] or MRI) and/or core infarct volume ≥50 mL (on CT perfusion [CTP]: rCBF <30%);
6. Occlusion of the intracranial internal carotid artery (ICA), the M1- or M2-segment of the middle cerebral artery (MCA) confirmed by CT angiography (CTA), MR angiography (MRA), or digital subtraction angiography (DSA);
7. Planned treatment with EVT;
8. Informed consent obtained from patients or their legal representatives.

##### Exclusion criteria

Patients meeting any of the following criteria will be excluded from study enrolment.

1. Intracranial hemorrhage confirmed by cranial CT or MRI;
2. mRS score >2 before onset;
3. Pregnant or lactating women;
4. Allergic to contrast agents or glucocorticoids;
5. Participating in other clinical trials;
6. The artery is tortuous such that the thrombectomy device cannot reach the target vessel;
7. Bleeding history (gastrointestinal and urinary tract bleeding) within 1 month;
8. Chronic hemodialysis and severe renal insufficiency (glomerular filtration rate < 30 mL/min or serum creatinine >220 umol/L [2.5 mg/ dL]);
9. Life expectancy due to any advanced disease <6 months;
10. Follow-up is not expected to be completed;
11. Intracranial aneurysm and arteriovenous malformation;
12. Brain tumor with imaging mass effect;
13. Systemic infectious disease.

### Randomization

Following eligibility confirmation and informed consent, patients will be centrally randomized in a 1:1 ratio to receive either intravenous methylprednisolone or placebo (normal saline). Randomisation will be performed immediately using a 24-hour real-time web-based system (via a WeChat Mini Program developed by Hefei Rongshi Information Technology Co., Ltd.), which employs a permuted block design to ensure balanced group allocation. The time of server confirmation of randomisation is defined as time zero for the study. Each enrolled patient will receive masked medications corresponding to their assigned random code based on their enrollment time. Both patients and trial personnel will remain blinded to treatment assignment throughout the study.

### Interventions

The methylprednisolone sodium succinate and its corresponding placebo (normal saline) used in this trial had the same formulation, production process, packaging, dosage, and administration as used in the MARVEL trial [1]. Each patient will receive intravenous methylprednisolone sodium succinate or its corresponding placebo of normal saline at a dose of 2 mg/kg (maximum 160 mg) once daily for 3 consecutive days, with the first dose initiated as soon as possible after randomisation.

### Imaging protocol

All imaging evaluations will be conducted under the supervision of a central imaging core lab, led and quality-controlled by international experts.

At baseline, site investigators will perform initial assessments of ASPECTS (based on NCCT or Diffusion-Weighted Imaging [DWI] MRI), infarct core volume (CTP, rCBF <30%), and arterial occlusion location (based on CTA/MRA/DSA). These imaging data are then transmitted to a central imaging core lab for real-time, independent review by two blinded clinicians, with a third senior reviewer resolving discrepancies. Enrollment requires an ASPECTS <6 and/or an infarct core volume ≥50 mL, plus confirmation of an intracranial ICA or M1/M2 of MCA occlusion.

Angiographic imaging, including the occlusion site and expanded Thrombolysis in Cerebral Infarction (eTICI) scores before and after thrombectomy, will be independently assessed by members of a central imaging core lab.

Follow-up imaging (NCCT/MRI within 48 hours) will be evaluated for hemorrhagic events by the imaging core lab. The infarct volume and quantitative edema biomarkers (midline shift, relative hemispheric volume, and net water uptake) [21, 22] will be automatically extracted from CTP, and NCCT or MRI using commercial F-stroke software (Version 1.0.21, Neuroblem ltd. Company, Shanghai, China), with manual adjustment as needed.

### Study procedures and assessments

Eligible patients presenting within 12 hours of last known well will provide informed consent prior to enrollment (Visit 0), during which baseline demographic, clinical, and imaging data will be collected. After confirmation of eligibility, patients will be randomized to receive methylprednisolone or placebo. The study drug will be administered intravenously once daily for three days, starting immediately post-randomisation (Visit 1, 12–48 hours). During this period, patients will undergo EVT and be monitored for vital signs, neurological status (assessed by NIHSS), safety outcomes, and imaging findings by NCCT/MRI. Follow-up assessments will be conducted at 5–7 days or discharge (Visit 2) for neurological status (assessed by NIHSS) and at 90 days for survival status and functional outcomes (assessed by mRS and European Quality Five-Dimension Five-Level Visual Analogue Scale [EQ-5D-5L VAS]), alongside safety evaluations. Long-term functional outcomes (assessed by mRS and EQ-5D-5L VAS) will be assessed at 1 and 5 years as tertiary endpoints.

### Efficacy endpoints

The primary efficacy endpoint is all-cause mortality at 90 (±14) days.

Secondary efficacy endpoints include:

1. Time from randomisation to the occurrence of death from any cause at 90 (±14) days;
2. mRS score at 90 (±14) days (scores 5 and 6 are merged);
3. Proportion of patients with mRS score 0 to 4 at 90 (±14) days (%);
4. Proportion of patients with mRS score 0 to 3 at 90 (±14) days (%);
5. Proportion of patients with mRS score 0 to 2 at 90 (±14) days (%);
6. Proportion of patients with mRS score 0 to 1 at 90 (±14) days (%) or return to pre-stroke mRS score (if pre-stroke mRS > 1);
7. Midline shift at 48 hours;
8. Proportion of patients with midline shift maximum >5 mm within 48 hours (%);
9. Relative hemispheric volume at 48 hours;
10. Net water uptake at 48 hours;
11. Proportion of patients with decompressive craniectomy after EVT (%);
12. NIHSS score at 5-7 days or at early discharge;
13. Health-related quality of life (EQ-5D-5L VAS) at 90 (±14) days.

Tertiary efficacy endpoints include:

1. mRS score at 1 year (scores 5 and 6 are merged);
2. Proportion of patients with mRS score 0 to 2 at 1 year (%);
3. Proportion of patients with mRS score 0 to 1 at 1 year (%) or return to pre-stroke mRS score (if pre-stroke mRS > 1);
4. mRS score at 5 years (scores 5 and 6 are merged);
5. Proportion of patients with mRS score 0 to 2 at 5 years (%);
6. Proportion of patients with mRS score 0 to 1 at 5 years (%) or return to pre-stroke mRS score (if pre-stroke mRS > 1);
7. EQ-5D-5L VAS at 1 year;
8. EQ-5D-5L VAS at 5 years.

### Safety endpoints

The primary safety endpoint is the proportion of patients with SICH within 48 hours after EVT (based on the modified Heidelberg Bleeding Classification [HBC]) [23]

The secondary safety endpoints include:

1. Proportion of patients with any ICH within 48 hours after EVT (based on modified HBC) [23];
2. Proportion of patients with pneumonia (%) [24];
3. Proportion of patients with gastrointestinal haemorrhage within 7 days after EVT (%);
4. Incidence of any complications;
5. Incidence of any (serious) adverse events.

### Data safety and monitoring board

An independent Data and Safety Monitoring Board (DSMB), composed of an experienced neurologist, a neuroradiologist, and a biostatistician, will have meetings annually and oversee patient safety and trial conduct throughout all phases of MIRACLE. All DSMB members must have no conflict of interest regarding the trial’s conduct. A charter detailing the DSMB’s membership, roles, and responsibilities will be formally ratified by both the DSMB and executive committee prior to enrollment. Following each meeting, the DSMB chair will promptly communicate the boards’ recommendations to the steering committee.

### Sample size

The current trial is primarily designed to investigate all-cause mortality in AIS patients with large infarct core within 12 hours of the last known well time who undergo EVT combined with early administration of methylprednisolone. Preliminary 90-day mortality data for AIS patients with large infarct core (based on ASPECTS <6) from subgroup analysis were obtained by contacting the corresponding author of the MARVEL trial [1]. Based on subgroup analysis data from the MARVEL trial and SELECT2 trial [5], and incorporating the clinical experience of neurointerventional neurologists, the expected proportion of mortality was 38% in the placebo group. We conservatively estimated a 9% decrease in mortality in the methylprednisolone group. Hence, we expected mortality would be 38% in the placebo group and 29% in the methylprednisolone group. The sample size ratio between the two groups was 1:1. Assuming a two-sided α = 0.05 and a power of 80%, the estimated sample size was 856 patients. Considering a 5% dropout rate, the total sample size was calculated to be 902 patients using PASS 15.0, with 451 patients in each of the methylprednisolone and placebo groups.

### Statistical analyses

The primary analysis will follow the intention-to-treat principle. The per-protocol analyses will also be performed as supplemental analyses. The primary outcome, all-cause mortality at 90 days, will be compared between groups using a modified Poisson regression model to calculate risk ratios (RR) with 95% confidence intervals (CI). Secondary outcomes will be analysed using appropriate statistical methods: Cox regression for time-to-death, win ratio for mRS shift analysis [25], modified Poisson regression model for binary secondary outcomes (such as mRS 0-2), and generalized linear models or win ratio for continuous outcomes (such as midline shift). Analyses will be adjusted for key prognostic covariates, which include age, baseline NIHSS score, pre-stroke mRS score, baseline ASPECTS, the time from last known well to randomisation, and occlusion site. Every effort will be made to minimize missing data through active follow-up and mortality ascertainment via local committees. The primary efficacy analysis will use a complete-case approach, with sensitivity analyses conducted to assess the robustness of results under different missing data assumptions. Missing key prognostic covariates will be handled using simple random imputation for variables with <5% missing data, and multiple imputation for those with ≥5% missing data. Pre-specified subgroup analyses will be conducted by age (median), sex, baseline NIHSS score (median), baseline ASPECTS (0-2 vs. 3-5), time from last known well to randomisation (median), intravenous thrombolysis (yes vs. no), occlusion location (ICA vs. MCA), infarct core volume (median), and reperfusion (eTICI grade 0-2a vs. 2b-3). A detailed statistical analysis plan will be finalized before database lock. All statistical analyses will be performed using SAS^®^ Software version 9.4 in a Windows environment and R version 4.1.1. Two sided with *p* <0.05 will be considered significant.

## DISCUSSION

The MIRACLE trial is designed to provide evidence on whether early adjunctive administration of intravenous methylprednisolone can improve survival and functional outcomes in AIS patients with large infarct core undergoing EVT within 12 hours of last known well time. This trial directly addresses the critical knowledge gap left by the MARVEL trial, which, while demonstrating a potential safety signal and reduced mortality in a broad LVO population, excluded the highest-risk patients with the largest infarcts (ASPECTS <3 or core volume >50–70 mL) [1, 26]. These patients are precisely the ones most vulnerable to life-threatening cerebral edema and hemorrhagic transformation—the very complications that corticosteroids are theorized to mitigate.

The current therapeutic cornerstone for AIS focuses on early restoration of blood flow and salvage of the ischemic penumbra. For patients with large infarct core due to anterior circulation LVO, EVT has been established as an effective standard of care, achieving high recanalization rates of 80-94% [9]. Yet, a significant proportion of patients with large infarct core experience poor outcomes or death [26]. Large core infarcts represent a state of extensive ischemic necrosis, which triggers a potent inflammatory cascade upon reperfusion. The disruption of the blood-brain barrier, activation of glial cells, and infiltration of leukocytes contribute to vasogenic and cytotoxic edema, leading to increased intracranial pressure and secondary neurological deterioration [12, 27]. In particular, large infarct core patients with ASPECTS 3-5 and/or infarct volumes >50-70 mL are at a markedly elevated risk of decompressive craniectomy, approaching 10% in the Endovascular Therapy for Acute Ischemic Stroke with Large Infarct (ANGEL-ASPECT), The Efficacy and Safety of Thrombectomy in Stroke with extended lesion and extended time window (TENSION), and Endovascular Therapy for Acute Stroke

With a Large Ischemic Region (RESCUE-Japan LIMIT) trials [3, 4, 6], as well as up to ∼22% in the trial of Thrombectomy for Stroke With Large Infarct on Noncontrast CT (TESLA) [8]. Moreover, in patients with large infarct core and unsuccessful EVT recanalization, the ANGEL ASPECT trial showed that there was greater infarct growth and higher rates for hemicraniectomy (19%) compared to patients who were medically managed [28]. The mortality rates of large infarct core patients reached up to 40% [5].

This discrepancy between high rates of successful recanalization and poor outcomes or death, is often attributed to cerebral edema, a central pathological driver of neurological deterioration that peaks within days of stroke onset. Recent evidence from the ANGEL-ASPECT trial indicates that EVT itself may induce an early increase in midline shift—a marker of mass effect—within 24 hours post-procedure. This early mass effect partially mediates poorer functional outcomes, despite the overall benefit of thrombectomy [21]. Moreover, quantitative neuroimaging studies have demonstrated that edema reduction, measured by net water uptake, mediates up to 66% of the treatment effect of recanalization on functional outcome. This contribution is substantially greater than the effect attributable to penumbra salvage alone. [14] These findings underscore the critical role of edema in limiting the full potential of reperfusion therapies and highlight the need for adjunctive neuroprotective strategies targeting edema formation.

Inflammatory mechanisms are promising therapeutic targets in stroke, with several proof-of-concept trials already demonstrating the efficacy of immune modulators as an adjunctive therapy to EVT. Preclinical and early-phase trials have suggested potential neuroprotective effects of agents like fingolimod, butylphthalide, and Aptoll when combined with EVT [29-31]. However, the high cost and limited availability of such targeted therapies currently restrict their widespread clinical application. Corticosteroids, with their potent anti-inflammatory, immunomodulatory, and blood-brain barrier-stabilizing properties, represent a mechanistically plausible intervention to disrupt this deleterious process [17]. Preclinical evidence robustly supports that early corticosteroid administration can reduce brain edema and limit infarct expansion [18, 32, 33]. The exploratory findings from MARVEL suggest this translational potential may extend to humans with ASPECTS 3–10 [1]. However, the exclusion of the most severe cases of EVT patients left a critical evidence gap.

MIRACLE is unique among ongoing neuroprotection studies in its specific focus on a high-morbidity population with large ischemic core (ASPECTS <6 and/or core volume ≥50 mL), where the balance between benefit and risk may be most favorable if edema mitigation can be achieved. In addition, MIRACLE designs several elements to rigorously assess the impact of methylprednisolone on cerebral edema. These include quantitative imaging biomarkers such as midline shift, relative hemispheric volume, and net water uptake at 48 hours—objective measures that directly reflect edema severity [21, 22]. By including these secondary endpoints, the trial aims to provide mechanistic insights into how methylprednisolone may exert its effects.

Nevertheless, potential limitations must be acknowledged. First, the trial is conducted exclusively in China, which may limit the generalizability of the findings to other ethnic populations with potentially different stroke characteristics or comorbidities. Future studies may be needed to validate the results in other ethnic populations. Second, this study included only patients with anterior large vessel occlusion, and patients with acute basilar occlusions should be further studied.

## CONCLUSION

In summary, MIRACLE is a pathophysiologically based, pragmatically designed trial that targets a high-risk population with large infarct core where cerebral edema could be a dominant driver of a patient’s outcome. By evaluating both clinical and imaging endpoints, it seeks to determine whether early anti-inflammatory therapy with methylprednisolone can reduce mortality and improve functional outcomes after successful thrombectomy, potentially establishing a new standard of adjunctive care for this devastating stroke subtype.

## Data Availability

Data will be available for reasonable reques from.

## Acknowledgements

We thank all the participants in the MIRACLE program for their hard work.

## Ethics approval

This study involves human participants and all protocol modifications were submitted and approved by ethics committee at First Affiliated Hospital of Fujian Medical University (MRCTA, ECFAH of FMU [2024]368-1) and all participating centers. The study is conducted in accordance with Good Clinical Practice and the Declaration of Helsinki. All patients or their legal representatives must provide written consent.

## Data availability statement

Data are available upon reasonable request

